# Community Engagement to support COVID-19 Vaccine Uptake: A living systematic review protocol

**DOI:** 10.1101/2022.03.08.22272082

**Authors:** Brynne Gilmore, Nina Gerlach, Claudia Abreu Lopes, Alpha Ahmadou Diallo, Sanghita Bhattacharyya, Vergil de Claro, Rawlance Ndejjo, Elizabeth Nyamupachitu- Mago, Adalbert Tchetchia

## Abstract

**Background:** Widespread vaccination against Coronavirus Disease 2019 (COVID-19) is one of the most effective ways to control, and ideally, end the global COVID-19 pandemic. Vaccine hesitancy and vaccine rates vary widely across countries and populations and are influenced by complex socio-cultural, political, economic, and psychological factors. Community engagement is an integral strategy within immunisation campaigns and has been shown to improve vaccine acceptance. As evidence on community engagement to support COVID-19 vaccine uptake is emerging, this review aims to lessen the knowledge-to-practice gap by providing regular evidence on current best-practice.

**Methodology:** A living systematic review will be conducted which includes an initial systematic review and bi-monthly review updates. Searching and screening for the review and subsequent updates will be done in four streams: a systematic search of six databases, grey literature review, preprint review, and citizen sourcing. All articles will be collated into Covidence, where screening will be done by a minimum of two reviewers at title/abstract and full-text. Data will be extracted across pre-defined data extraction tables, and synthesis will occur using the convergent integrated approach. Updates to the review resulting from the subsequent bi-monthly searches will be shared in an open-access platform. The protocol has been registered with PROSPERO: CRD42022301996.

**Discussion:** Given the variation in vaccination rates across different contexts and the recognition that high vaccination coverage is required to reduce COVID-19 transmission and to stop the emergence of new variants, it is imperative the global community implements strategies that will improve uptake and that this work is widely shared and contextualised. Community engagement to promote vaccine uptake is highly encouraged, and recent studies highlight its potential to influence vaccine rates, particularly across populations that are marginalised. The high-priority research needed on this topic, and the rapidly changing evidence base, supports the conduct of a living systematic review.

## Introduction

### COVID-19 Vaccine uptake and hesitancy

Widespread vaccination against Coronavirus Disease 2019 (COVID-19) is one of the most effective ways to control, and ideally, to end the global COVID-19 pandemic. Even when vaccine supply is available and consistent, differences in rates of vaccination uptake can be observed across countries and sub-populations. For instance, a recent systematic review that assessed COVID-19 vaccine acceptance rates found the highest acceptability in Ecuador, Malaysia, and Indonesia at 97%, 94.3%, and 93.3%, respectively (1). Among the general population, Kuwait indicated the lowest acceptance rate at 23.6%, followed by Jordan (28.4%) and Italy (53.7%) (1). The updated version of the same review found that in East and Southern Africa (n = 9), the highest COVID-19 vaccine acceptance rate was in Ethiopia (92%) and the lowest in Zimbabwe (50%). In West/Central Africa (n = 13), Niger (93%) had the highest rate, and Cameroon (15%) the lowest (2). Among health workers, Israel reported the highest COVID-19 vaccine acceptance rate at 78.1%, while a survey from the Democratic Republic of the Congo indicated a rate of 27.7% (1). However, results across countries are not directly comparable as different survey methods are used. For example, the sampling strategies, response rates, and mode of administration (online, telephone, face to face) vary widely across these national surveys.

Vaccine hesitancy is defined as the ‘delay in acceptance or refusal of vaccination despite the availability of vaccination services’ (3). This term refers to a continuum and encompasses a heterogeneous group of individuals who range between those who clearly accept all vaccines and those who undoubtedly decline all. Vaccine hesitancy is a complex phenomenon as it is underpinned by a mix of economic, psychological, socio-cultural, and political factors (4).

The World Health Organization (WHO)’s Strategic Advisory Group of Experts on Immunization (SAGE) identified three key reasons for vaccine hesitancy: confidence, convenience, and complacency – called the “3Cs” model. Confidence refers to the trust that a vaccine is safe and effective, as well as trust in the health system and in the motivations of policymakers. Convenience is defined by vaccine affordability, availability, geographical accessibility, and health literacy. Complacency refers to the perception that the disease risk is low and receiving the vaccine is not needed (3).

In line with the 3Cs model, specific reasons related to COVID-19 vaccine hesitancy cited in the literature include perceptions of vaccine efficacy, effectiveness and safety, worries about the side effects, confidence and preference for domestically made vaccines, political values and context, conspiracy theories, and anti-vaccination rumours and misinformation from social media platforms (5,6). In targeted groups like health care workers or minorities, some additional factors include trusting the immune system to combat the virus, insufficient knowledge about vaccines, and politics surrounding vaccine development processes (7,8). Specific to minorities, socio-economic characteristics, perceived risk and convenience in obtaining the vaccines have been reported (9).

Vaccine hesitancy is exacerbated by a lack of health literacy and also misinformation from social media and other information channels (10,11). During the pandemic, the rapid increase in the volume of information created an ‘infodemic crisis’ (12). The term infodemic refers to an exponential increase in the volume of information associated with a global issue which also includes misinformation (false information unintentionally shared or spread) and disinformation (false or inaccurate information deliberately intended to deceive).

Given that the factors that lead to vaccine hesitancy vary across socio-demographic groups, social stratifiers such as gender, age, race, education level, marital and economic status are also associated with vaccine acceptance (9,13). Despite mixed evidence, some research has shown that COVID-19 vaccine acceptance varies across ethnic groups (14). In the United Kingdom (UK), four in ten adults of Black or Black British heritage are COVID-19 vaccine-hesitant, compared to one in ten White British adults (15). Being a woman is associated with a greater hesitancy towards COVID-19 vaccines (16,17). However, a systematic review commissioned by SAGE found that education and socio-economic status did not affect vaccine uptake in the UK. A higher level of education could be linked to both an increased, as well as a decreased acceptance (18).

### Community engagement for vaccine uptake in COVID-19

Community engagement has been part of global recommendations and guidelines on the response to the COVID-19 pandemic. However, its implementation has not always been effective (19), though lessons learned from previous vaccination programmes show that community engagement is an effective and essential tool (20). In South Asia, for example, community engagement efforts in Ebola and polio vaccines were used consistently and successfully (21). A recent review on community engagement for prevention and control of infectious disease (19) noted how community engagement has been used to support vaccine uptake and recommends using such efforts for COVID-19. Community engagement can be used as community entry plans, for co-designing vaccination strategies and messaging, to disseminate timely information on vaccines and immunisation strategies, and to build trust and address misinformation (22).

SAGE identifies enabling environments among drivers for COVID-19 vaccine acceptance and uptake. Among those, social norms conducive to vaccination can be created and reinforced in groups by the community, religious leaders, and civil society organisations (23). This implies that communities lead on issues that affect them to use vaccination services and build resilience (24). Another publication both of WHO and the United Nations International Children’s Emergency Fund (UNICEF) point out the specific roles of community health workers (CHWs) in COVID-19 vaccination to support the buy-in and uptake of vaccination from communities and individuals (25).

Evidence on community engagement for COVID-19 vaccine uptake is emerging (26). For example, it has been successfully used to increase Black, Indigenous and People of Colour’s participation in COVID-19 clinical trials in the USA and to increase vaccine compliance among Arab and ultra-orthodox Jewish populations in Israel (27,28). Good examples of citizen engagement from Malaysia involve community leaders reaching out to the indigenous population and the United Nations High Commissioner for Refugees’ (UNHCR’s) efforts to reach undocumented migrant workers and refugees (29,30). UNICEF established the U-report information chatbot to support COVID-19 risk communication and community engagement in 52 countries among youth and communities (31). In Pakistan, 13,000 female-led teams of health workers went door to door in Sindh province to campaign and offer vaccines to 25% of the population who had not received any dose, mainly traditional women who are less literate (32).

Evaluation of COVID-19 vaccination rollout has also shown that community engagement came out of measures to mitigate major challenges threatening the success of the vaccination in Africa (33). Given the newly developed vaccines and their increase in availability, the emerging COVID-19 variants, the need for boosters, and the urgency to end the pandemic, the relevance of community engagement remains critical and thus the urgent need for evidence to inform current efforts. However, there is a dearth of evidence on how community engagement can be used to support vaccine uptake (34). Collating the emerging and evolving evidence base on community engagement to support COVID-19 vaccine uptake is therefore required which is the gap that this review aims to bridge.

## Methodology

### A living systematic review

A living systematic review (LSR) is a systematic review that is continually updated according to an explicit a priori schedule (35). They are not themselves a review methodology, but an approach to updating reviews (35). Relevant new evidence for the review is incorporated as it arises, as the process supports the continual and active monitoring of evidence (36). As such, a LSR aims to provide readers with a single source to review up-to-date, high-quality evidence on a specific topic (37). Living systematic reviews are ideal for situations when the field and evidence is rapidly developing with new evidence emerging (38) and for high priority topics (39). The importance of living evidence has been outlined and advocated for within COVID-19 to support the rapidly evolving evidence-based approach to address the ‘knowledge to practice’ gap (40).

Undertaking an LSRs is consistent with other systematic review methodologies, with key features of LSRs including: specification of how frequently new evidence is searched for when evidence is incorporated into the review (36), and having online-only evidence summaries that are frequently updated (41).

LSRs have the potential to reduce workload by working off existing efforts and streamlining the research approach, and avoiding research duplication (37). Challenges to conducting LSRs include the human resource commitment (37), lack of methodological tools, such as data management programmes that are tailored to LSRs (41), and little to no guidance for reporting LSRs (39,42). Recommendations for the conduct of LSRs include exploring the use of ‘citizen science’, participation such as crowdsourcing (41), and ensuring transparent reporting of review methodology and updates (39). In the context of rapidly emerging evidence, such as the case with COVID-19, it may be necessary to allow for reviewing and updating of methodological processes, including post-hoc changes to inclusion criteria, to include preprints as sources and a variety of searching methods (39).

There are numerous ongoing LSRs on COVID-19 topics including clinical trial registration (43), drug treatments (44), characterising long COVID (45) and mental health outcomes (46). Given the global magnitude of COVID-19 and the rapidly changing evidence environment, an LSR is an appropriate methodology for the high priority topic of community engagement activities used for vaccine uptake.

### Review Objectives and Questions

This review aims to lessen the knowledge-to-practice gap for using community engagement to support vaccine uptake for COVID-19 by providing regular evidence on current best-practice. To do so, it will:

1. Conduct a rigorous systematic review on community engagement for COVID-19 vaccine uptake.
2. Update the review on a bi-monthly basis using set procedures.
3. Disseminate updated findings and recommendations on an open-access platform.

To this end, it will endeavour to answer the following research questions by conducting an LSR:

1. How is community engagement being used to support COVID-19 vaccine uptake and/or reduce hesitancy, including the characterization of different components of the community engagement process?
2. What is the effect of this engagement on vaccine uptake and/or reduction in vaccine hesitancy?
3. What implementation lessons for using community engagement for vaccine uptake can be learned, and how do these differ across population groups and settings?

### Methods and Tools

The protocol for this review is divided into two phases. First, the initial systematic review procedures, including searching, inclusion/exclusion criteria and data extraction will be conducted. Second, updated searching procedures to make the review ‘living’. The protocol has been registered with PROSPERO: CRD42022301996.

Step 1: Initial Review

An initial systematic review to address the aforementioned research questions will be conducted. Given the limited time frame for searching (from January 2020) and our research team size, we anticipate this systematic review will take approximately two to three months to complete. The intention will be to publish this review, with links to the open-access platform where updates arising from the iterative bi-monthly searching, and any revisions to the methodology, will be shared.

The initial search is anticipated to begin in mid-March 2022, following the below methods.

#### Inclusion and exclusion

Articles will be included if they detail community engagement for improving vaccine uptake and/or reducing hesitancy for individuals eligible for COVID-19 vaccines. Articles must provide insight into either how community engagement has been used to support vaccine uptake, the effectiveness of community engagement for vaccine uptake, or both. Articles may report specifically on vaccine figures, or provide insights into how community engagement can work based on primary evidence. For instance, a qualitative article may report on community members’ experience with community engagement, but not highlight the percentage of uptake. Articles detailing community engagement efforts to support uptake prior to vaccine roll-out in the specific location will be included if they detail efforts to increase vaccine acceptability and/or reduce vaccine hesitancy. Searching will be done in English, however, no language restrictions will be applied. In table 1, inclusion and exclusion criteria are outlined in more detail.

**Table 1:** Inclusion and exclusion criteria.

| Topic | Inclusion | Exclusion |
| --- | --- | --- |
| **Population – vaccine eligible** | Any individual, regardless of age, eligible for COVID-19 vaccines.<br>Any government approved and supported COVID-19 vaccine: including initial dose and subsequent doses (booster etc.). | Individuals receiving vaccines as part of clinical trials.<br>Engagement of parent/guardian for vaccination in children. |
| **Exposure – community engagement** | Community engagement activity to support vaccine uptake and/or reduce hesitancy. | Not community engagement, or community engagement focus not to increase vaccination acceptance or uptake. |
|  | What is community engagement in this review: An approach that involves inclusion and participation of individuals, groups or structures within the parameter of a social boundary or catchment area ('the community') to influence a health outcome or behaviour, or to support community decision-making, planning, design, governance and delivery of service (modified definition from (47)).<br>What is not community engagement in this review: One-way communication efforts targeted at communities and/or individuals such as: media (radio, tv, social media) campaigns; information distribution including pamphlets, mail; and counselling by health worker, including door-to-door by CHWs, or consultation by healthcare provider. |  |
| **Outcomes** | Articles will be included if they address the primary and/or secondary outcomes below:<br>**Primary outcomes:**<br>• Vaccination uptake<br>• Vaccine acceptability/intention to vaccinate<br>• Vaccine hesitancy<br>**Secondary outcomes:**<br>• Implementation considerations for using community engagement for vaccines<br>• Insights into how community engagement can support vaccines<br>• Knowledge and awareness about vaccines<br>• Attitudes towards COVID-19 vaccines | Reports community engagement used but not enough information to extract insight |
| **Location** | Worldwide | No restrictions |
| **Timeframe** | January 01, 2020 to present<br>Note: articles will be included if they have community engagement to support vaccines prior to vaccine administration if they meet any of the above criteria on outcomes | Pre-January 01, 2020 |
| **Article Type** | Primary research, both qualitative and quantitative and all study designs. Preprints included. | Not primary research, including opinions, commentaries and guidelines. Secondary research, including reviews. |

#### Searching for articles

To support a robust searching process on this rapidly developing topic, we will utilise four searching techniques: database search, preprint search, grey literature search, and citizen sourcing. The following databases will be searched: PubMed, CINAHL, Embase, Cochrane Library, LILACS and AJOL. Grey Literature will be searched via <u>WHO’s COVID-19 Research Database</u>. Given the rapidly evolving evidence base for COVID-19, research publication may lag behind research completion. As such, we will use both preprints and citizen sourcing to identify completed activities that have yet to be catalogued or are under review/revision. The health science preprint servers medRxiv and bioRxiv will be searched. Citizen sourcing will involve the creation of a Twitter and email account. These will be used to identify other relevant groups, disseminate the search topic and solicit resources, and for the email account email relevant listservs. All included articles’ references will also be hand searched.

Three search topics will be used with a combination of MESH and Boolean phrases for the database search: Vaccine, COVID-19 and community engagement. Topics will be combined by ‘AND’. Table 2 provides example terms.

**Table 2:**
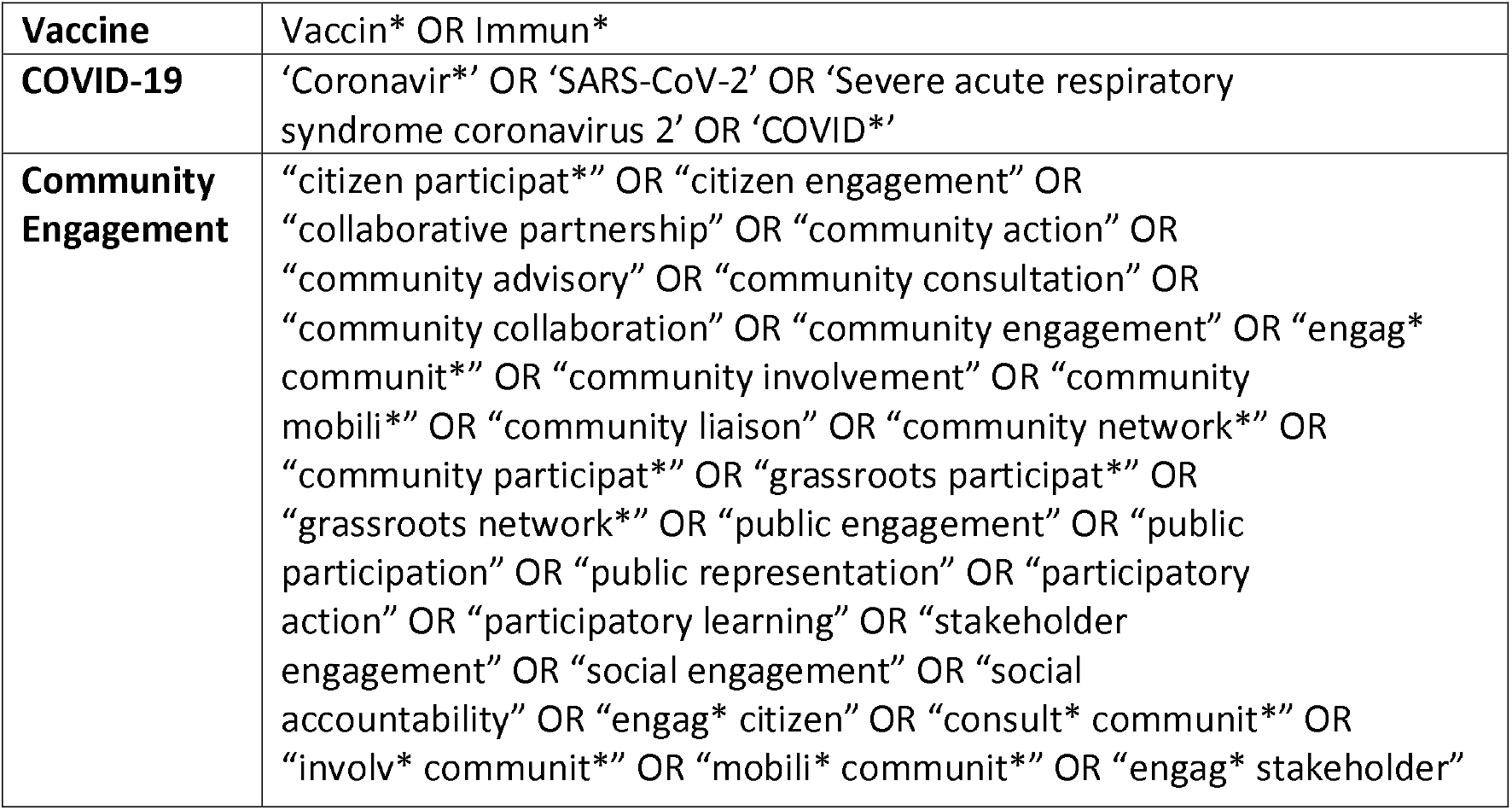
Example search terms.

#### Screening

After searching is complete, all returned references from the four techniques will be added to Covidence systematic review management software. Duplicates will be identified and removed, after which title/abstract screening will be conducted by two authors independently. Any discrepancies will be handled by a third reviewer. Full-text will be retrieved and screening will occur by two authors independently, with discrepancies again managed by a third author. A third reviewer will also randomly review 20% of articles screened at full-text stage for additional interrater reliability. All authors will review the list of articles at this stage, and consensus amongst the research team on the final included articles will be sought.

#### Data extraction and synthesis

All included articles will undergo data extraction by using a pre-defined data extraction template. Data related to article characteristics, context and population, and community engagement approaches and outcomes will be extracted, as highlighted in table 3. Extraction will occur independently by two reviewers, with findings compared and consolidated. Any discrepancies in extraction will be discussed with a third reviewer.

**Table 3:** Data extraction template.

| Characteristics<br>Context and<br>Population | Author/Date | Location | Vaccine rates<br>(pre-community<br>engagement) | Vaccine<br>Commentary* | Target<br>population | Population<br>characteristics*<br>* | MMAT |
| --- | --- | --- | --- | --- | --- | --- | --- |
| Community<br>Engagement | Definition of<br>community<br>engagement | Community<br>engagement<br>description,<br>justification | Who did<br>community<br>engagement:<br>Employment,<br>Training, Payment | What did they<br>do | When, where<br>and how<br>often | Community<br>engagement<br>reported<br>outcomes | Equity<br>considerations |
|  | Implementation<br>Characteristics | Individual Level:<br>Inner setting:<br>Outer Setting: |  |  |  |  |  |
|  | Implementation<br>Evaluation and<br>barriers | Reach:<br>Efficacy:<br>Adoption:<br>Implementation:<br>Maintenance |  |  |  |  |  |

\*any reports on population/context vaccine views, including hesitancy, trust and confidence.
\*\*including residence location, gender, socio-economic status, ethnicity, resident status etc.

The convergent integrated approach for mixed methods systematic reviews (48) will be used to synthesise findings for research questions one and three. Using the data extraction tables, this will involve the key concepts of data transformation (i.e. ‘qualitizing’ any quantitative evidence to have comparable data). After qualitizing, integration will occur following the principles of meta-aggregation (49). Categories of similar findings from the studies will be identified, and then synthesised into the final review findings. The initial categories will be identified by first becoming familiar with the data through repeated reading of the extraction tables, and second by having two authors independently propose categories. Authors will then discuss categories and share the proposed categories, with detailed examples, with the remainder of the research team to gain consensus. Once the categories are established, all data will be re-reviewed and coded to relevant categories. Synthesis of categories into overall study findings will follow a similar process.

For research question two, given the diversity of study and intervention approaches that can be taken within community engagement, it is unlikely that a meta-analysis will be possible. However, we will explore the potential to quantitatively synthesise any primary outcomes (vaccine uptake, vaccine acceptability, vaccine hesitancy) depending on heterogeneity of the studies. This will involve using a descriptive statistical approach. If the data allows, the team will undertake a pooled analysis using risk-ratio/odds-ratio (dichotomous outcomes) or mean difference (continuous outcomes).

A key findings summary table will be developed which will seek to answer the research questions and highlight key considerations for community engagement for vaccine uptake, including relevant references.

#### Risk of Bias

Given the article types that are included, the Mixed Methods Appraisal Tool (MMAT) will be used on all included articles. The MMAT score for all included articles will be recorded.

Step 2: Updating the Review

#### Searching

Bi-monthly database, grey literature and preprint server searches will occur. Dates will be adjusted to reflect the last search date, as to only return sources published within the previous two months. Citizen searching will be an ongoing process. Any returned resources from the four searching sources will be uploaded into Covidence to undergo screening at title/abstract and full-text phases by two research team members independently, consistent with the screening approach from the first step of the review. Discrepancies will be managed by a third reviewer.

It is anticipated that adjustments to the search terms and/or strategy may be required throughout the life cycle of this review. In these instances, changes will be clearly documented within the open-access protocol with a justification for the adjustment provided. If any changes to the search strategy reflect substantial variation from the initial protocol (as deemed by the research team) updated searching for those specific changes will be conducted.

Full prints of any preprints included in previous rounds of reviews will be sought. When a preprint is included, we will email the corresponding author to ask to be informed of any research or publication updates. If/when an included preprint becomes published, the previous preprint source will be updated to reflect the finalised article.

#### Data extraction and synthesis

After each bi-monthly iterative search and article screening, included articles will be added to the data extraction table. Extraction will be completed by one reviewer, with a second reviewer reviewing the completed extraction. Synthesis will occur by reviewing new data against the preceding data and highlighting similarities and differences. A quorum of the research team (minimum 4 members) will meet virtually to discuss the findings in light of any newly arising data and will interpret what this means for existing findings and recommendations. The key findings table will be updated accordingly.

It is anticipated that the organisation of the data extraction table and/or summary findings table will be revised as the review develops. The team anticipates that findings may be grouped along with geographical or contextual classifications, community engagement activity types, or target populations.

Step 3: Sharing review findings and updates

The initial review and subsequent bi-monthly searches and their results will be disseminated transparently and via open-access methods. A ‘read only’ GoogleDoc will be created, with links available in the published review from step one. Within this, main resources will be available: detailed protocol including any previous versions if revisions made, updated PRISMA flowchart with accompanying details of search dates and returns, the ‘living’ data extraction sheet including references for all included articles, key findings table, key programme recommendations, key recommendations for future research, and a recent updates table where changes made over the last two iterations will be featured.

As well as a continual open-access space for updating results, it is envisioned that a yearly open-access peer-reviewed publication will be developed. However, if emerging data strongly changes findings from the previous publication, we will endeavour to disseminate findings immediately. Quarterly briefs will be developed and shared on the reviews’ media platforms and across other interested networks and repositories. A dedicated web link will be created in the <u>Community Health-Community of Practice</u> site for sharing findings and also for obtaining feedback through webinars.

Social media accounts for the review, including email and Twitter accounts (@CE4_vaccines) will support searching for evidence and dissemination efforts. A mailing list will be developed, with interested parties being able to subscribe to any updates. Updates will be shared on Twitter as they arise, with regular posts seeking information on any new literature, ongoing or completed research that has emerged. Whereas all review files on GoogleDocs will be ‘read only’, we will have an additional page for readers to provide any additional resources, and comment on review findings and interpretation, aiming to increase both the searching process and the rigour and trustworthiness of the review.

## Discussion

The development of COVID-19 vaccines has come with the need for public health guidance on how best to garner support for vaccination, including reducing hesitancy and improving vaccine literacy. Given the current vaccination rates across different contexts, and the recognition that high vaccination coverage is required to reduce COVID-19 transmission and stop the emergence of new variants, the global community should implement strategies that will improve uptake. The importance of this topic is only to increase with the need for booster vaccines and new immunisation protocols to accommodate variants. As such, asking the question of how community engagement can, and is, being used to support vaccine uptake is crucial to advance this field and support COVID-19 vaccination efforts worldwide.

The high priority research needed on this topic, and the rapidly changing evidence base, supports the conduct of a LSR (40), which have gained traction owing to the COVID-19 pandemic. While caution on their conduct needs to be applied, especially on issues of data management and reporting, our review protocol has attempted to control for such potential limitations and learn from previous reviews. Specifically, we have included a preprint search and citizen searching, and will leverage social media via Twitter and GoogleDocs for identification of articles and feedback from the global community on the process and findings.

There are, however, recognized potential limitations of this review. Firstly, there are no specific systematic review software programmes that support monthly iterative searching, resulting in foreseeable challenges in data management. To limit any effects of this, all searches and their results will be saved and catalogued prior to uploading into Covidence. It is also anticipated that multiple data management systems may need to be used, and/or that flexibility across time will be required.

Secondly, conceptualizations of ‘community engagement’ will likely vary across settings. Such terminology is regular vernacular within some contexts, specifically the implementation of health programmes within low- and middle-income countries. Yet, even within this work, how community engagement is defined and what it encompasses is varied and often unclear (50,51). Moreover, the use of ‘community engagement’ as a term may be limited in contexts that implement fewer activities at the community level, for instance in high-income contexts or contexts with well-developed health systems. To reduce the influence of this potential challenge, the search terms and inclusion criteria aim to accommodate the many permutations of the term, as well as allow for flexibility and discretion for inclusion.

### What this review adds

This review adds to the global evidence base on public health interventions to control and prevent COVID-19. This protocol will allow for bi-monthly additions of new evidence and transparent and timely dissemination of review findings and updates on the important topic of community engagement for COVID-19. Policymakers, implementers, and researchers will be able to utilise this review to make evidence-based decisions from the most recent available data.

## Conclusion

The global burden of COVID-19 can only be substantially reduced with equitable access to vaccines and high vaccine uptake across all countries and populations. Community engagement is a required component of any successful vaccination campaign, and maybe especially crucial for populations that are marginalised. The current evidence base on how community engagement can support COVID-19 vaccination efforts is rapidly developing. Understanding how to best combat mistrust and build vaccine confidence, especially across different contexts and populations, will be required to improve vaccination uptake worldwide. Providing an up-to-date evidence repository on such efforts by conducting a LSR can enhance bridging the ‘knowledge-to-practice’ gap, and provide continuously evolving findings and key recommendations in line with the changing and crucial environment.

## Data Availability

All data produced in the living systematic review will be available in an open-access platform.

## Abbreviations

AJOL: African Journals OnLine
CHW: Community Health Worker
CINAHL: Cumulative Index to Nursing and Allied Health Literature
COVID-19: Coronavirus Disease 2019
LILACS: Latin American and Caribbean Health Sciences Literature
LSR: Living Systematic Review
MMAT: Mixed Methods Appraisal Tool
SAGE: Strategic Advisory Group of Experts on Immunisation
UNHCR: United Nations High Commissioner for Refugees
UNICEF: United Nations International Children’s Emergency Fund
WHO: World Health Organisation

## Authors note

The authors of this paper support global vaccine equity and highlight the need for equitable distribution of vaccines worldwide. All the authors of the paper except for NG are members of a collaborative platform, Community Health – Community of Practice.

## Availability of Review Data

Updates to the review and data in the form of data extraction sheets will be made available on the GoogleDrive. In addition, updates of the review will be publicly available on the Collectivity website (www.thecollectivity.org).

## Authors’ Contribution

All authors except NG were involved in a previous study on community engagement from which this current protocol took inspiration (18). BG conceived the study. All authors contributed to the study design development and manuscript writing.

## Public Involvement

The research team is made up of academics, policymakers, and implementers internationally. No additional public involvement measures are included.

